# Literature review on eye trauma in Africa

**DOI:** 10.1101/2025.01.16.25319949

**Authors:** Mapenzi Ndagonywa Philémon, Daniel Kakusu

**Author notes:** corresponding author (MNP): Faculty of Medicine, Evangelical University in Africa, Bukavu, DRC, (DK) Faculty of Medicine, Official University of Bukavu, Bukavu, DRC. These authors contributed equally to this work.

## Abstract

**Background:** Ocular trauma is a relevant cause of monoocular blindness in Africa. Although once neglected, it has recently been incriminated as one of the main etiologies of non-congenital visual impairment throughout Africa, the epidemiology of ocular injuries varies across regions, and remains an increasing concern, hence the interest in highlighting its aspects.

**Objective:** This study aims to take stock of the overall situation on the epidemio-clinical, etiological and therapeutic aspects.

**Methods:** This study is a narrative literature review of articles on ocular trauma in Africa published in Medline via Pubmed and Semantic scholar from Research rabbit. After cross-referencing the keywords, we found, relative to our study period from 2012 to 2022, 215 articles in the literature on eye trauma, of which nine articles were thus retained according to the inclusion and exclusion criteria.

**Results:** In relation to the frequency, the highest frequency according to the regions is 44.1% observed in the studies carried out in Tanzania in 2019 and the lowest is observed in the studies carried out in Nigeria in 2017 while two years later still in Nigeria the results showed an increasing trend to 30.7%; which proves that the frequency of eye trauma in Africa varies in time and space. Clinically, the results illustrate the most common clinical signs. It emerges that each study has its particularity on the most common signs. Whatever the spatio-temporal particularities of eye trauma, the most commonly used management is medical treatment.

## Introduction

Eye trauma is a generic term used to describe physical or chemical injury to the eye or orbit. **(1)** It is a lesion or a set of lesions resulting from the action of a physical agent or a chemical substance strictly affecting: the eyeball and its annexes, the orbit, and the optical pathways. **(2)** It can be defined as the set of morbid lesions of the eyeball due to external violence. **(3)**

The severity of eye injuries varies depending on the nature of the injuries. They are a major cause of visual impairment or loss of the eyeball. They constitute a public health problem. **(1,4)**

Occupational eye injuries are a global cause of visual morbidity. **(5)** According to WHO, 55 million cases of eye injuries occur each year; approximately 1.6 million lead to total blindness **(6)** and 2.3 million cause visual impairment, and another 19 million are cases of monocular blindness or visual impairment. **(7-8)**

Ocular trauma, once described as a neglected disorder, has recently been highlighted as one of the major etiologies of monocular, noncongenital visual impairment and blindness in all regions of the world. Ocular trauma accounts for 7% of all bodily injuries and 10% to 15% of all ocular diseases. **(9)**

Worldwide, there are approximately 1.6 million people who are blind due to eye damage, an additional 2.3 million people with bilateral low vision from this cause, and nearly 19 million with unilateral blindness or low vision **(10-12)**, which represents a significant economic burden due to the costs of treatment, hospitalization, family care, and time lost from work or school. **(12)**

Globally, ocular trauma accounts for 38–65% of all trauma cases in emergency departments and 5–16% of admissions to eye hospitals. **(13)**

Based on the incidence of eye injuries over the past decade worldwide, eye injuries remain a major cause of vision loss in developed, developing and high-income developing countries. **(14)**

In developing countries, eye injuries are not only more frequent but also more severe in their effects, which can be attributed to socio-economic background, inadequate safety measures, lack of optimal treatment facilities, use of traditional eye medicines and poor education. **(15)**

Particularly in developing countries, approximately half a million people worldwide are blind as a result of eye injuries and approximately 30–40% of monocular blindness is due to ocular trauma. The epidemiology of eye injuries varies across regions of the world, different age groups and depends on many factors including lifestyle, socioeconomic status, traffic conditions, sports and recreational activities and type of data recording. **(16)**

It is a common cause of emergency room visits, accounting for 36.3% of ophthalmological consultations in the United States, 26.1% of ophthalmological consultations in France and 3% of all consultations in Chile. **(17)**

The epidemiology surrounding ocular trauma has been reported in the United States, Europe, and Australia, but limited data on ocular trauma are available in China. Furthermore, although the incidence of ocular trauma in China may be higher than the incidences reported in some industrialized countries, the causes and types of injuries and prognosis have not been studied. **(18)**

In the United States alone, an estimated 2.0 to 2.4 million cases of ocular trauma occur each year, and nearly 1 million people suffer significant permanent visual impairment as a result of injury, with over 75% becoming monocularly blind. **(19)** In fact, it is estimated that as many as 27% of serious ocular injuries in the United States result in legal blindness (defined as visual acuity worse than 20/200). Trauma is consistently ranked as one of the leading causes of visual impairment in the industrialized world. **(20)** Ocular trauma accounts for 3% of all emergency department visits in the United States. **(21)**

In Brazil, approximately 705,200 occupational accidents were reported, of which 2,759 were classified as ICD (International Classification of Diseases) S05 (lesions of the eye and orbit) and 5,364 as ICD T15 (foreign body in the external eye). According to data provided by the Brazilian Social Security, the incidence of occupational accidents and ICD S05 decreased by 2.14% and 4.56%, respectively, while the incidence of ICD T15 increased by 11.59%, although these annual statistics do not include all work-related eye injuries but only accidents involving workers protected by labor legislation (CLT). **(22)**

In Korea, out of 1,900,000 cases, 70,000 (3.6%) cases of major trauma occur each year according to data collected by the Korean National Emergency Service Information System in 2018, and the incidence is increasing every year. **(23)**

In Australia, there are an estimated 20,000 hospital admissions due to eye injuries per year at a direct cost of $155 million. Indigenous Australians, men and those residing in rural areas have consistently been reported to be at higher risk of eye trauma. Despite the notable public health problem, recent population-based data on the frequency of serious eye trauma in Australian adults remain limited. **(24)**

A Finnish study on eye trauma showed that the incidence of eye injuries was 88/100,000/year. According to the Singapore Chinese Eye Study, eye injuries affect 1 in 25 adults; 20% of them required hospitalization, and men accounted for 74.6%. **(25)**

In Scotland, a study of children admitted to hospital with eye trauma found that blunt trauma accounted for 65% of injuries. **(26)**

In Spain, the percentage of patients with eye injuries sustained at work who receive urgent hospital care varies from 5.6% to 56.5%. In 2018, 17,579 workers were on sick leave due to a work-related eye accident. **(27)**

In Africa, they are a frequent reason for consultation in ophthalmology services. The frequency of these traumas varies according to the regions. It is around 12.4% in Northern Africa, 19.49 to 31.86% in Eastern Africa, 9.8% to 12.5% in Western Africa and 5% to 12.24% in Central Africa. **(2)**

In black Africa, the seriousness of traumatic eye emergencies (UOT) is increased by a lack of ophthalmologists and surgical equipment that is often unsuitable or even non-existent. **(28)**

In Senegal, eye trauma accounts for 12.5% of ophthalmological consultations. In Benin, it accounts for 10.4% of ophthalmology consultations. **(13)**

Although frequent in Morocco, these traumas have not been the subject of recent and targeted investigations. **(29)**

Studies in Nigeria and other parts of Africa have reported that ocular trauma is a significant cause of monocular blindness. **(15)**

In Guinea, studies have been conducted on eye trauma and have shown that the frequency of eye trauma in daily practice is high, concerning children who constitute a generally innocent vulnerable group and the complexity of its management had motivated the study. **(30)**

Although its frequency in Central Africa is around 12.24%, a search of published studies contrasts with the lack of data in this part, particularly in the DRC where there is really no data on eye trauma in the current literature. **(2, 31)**

In this study, we first need to comprehensively discuss the frequency of eye trauma, the age and/or gender most affected, the clinical signs and the most commonly encountered therapeutic methods.

## Methodology

### Type of study

The present study is a narrative literature review of articles on eye trauma in Africa published in Medline via Pubmed and Semantic scholar as well as from Research rabbit.

### Study population

This study concerns articles related to eye trauma in Africa.

### Inclusion criteria and non-inclusion criteria

#### Inclusion criteria

Only complete articles published in a peer-reviewed journal and archived online in the usage databases – Pubmed, Semantic Scholar and containing the following keywords were included in this study:

– Eye trauma,
– Eye injuries,
– Eye burns,
– Eye sores,
– Eyelid wounds,
– Africa,
– Only articles written in French or English were included in this study,
– Only open access articles were included in this study.

#### Non-inclusion criteria

We excluded these results obtained:

– Articles written in a language other than English or French,
– Those concerning black Africans from other continents,
– Incomplete articles, abstracts, scientific communications and blogs,
– Articles of the type commentary, response to the author and letter to the auditor, general report, review.

#### Sample size

After cross-referencing the keywords we found 215 including 15 duplicates, relative to our study period from 2012 to 2022, we found 97 articles in the literature on eye trauma.

#### Study period

This work is a review of articles on eye trauma in Africa over the last 10 years: from 2012-2022.

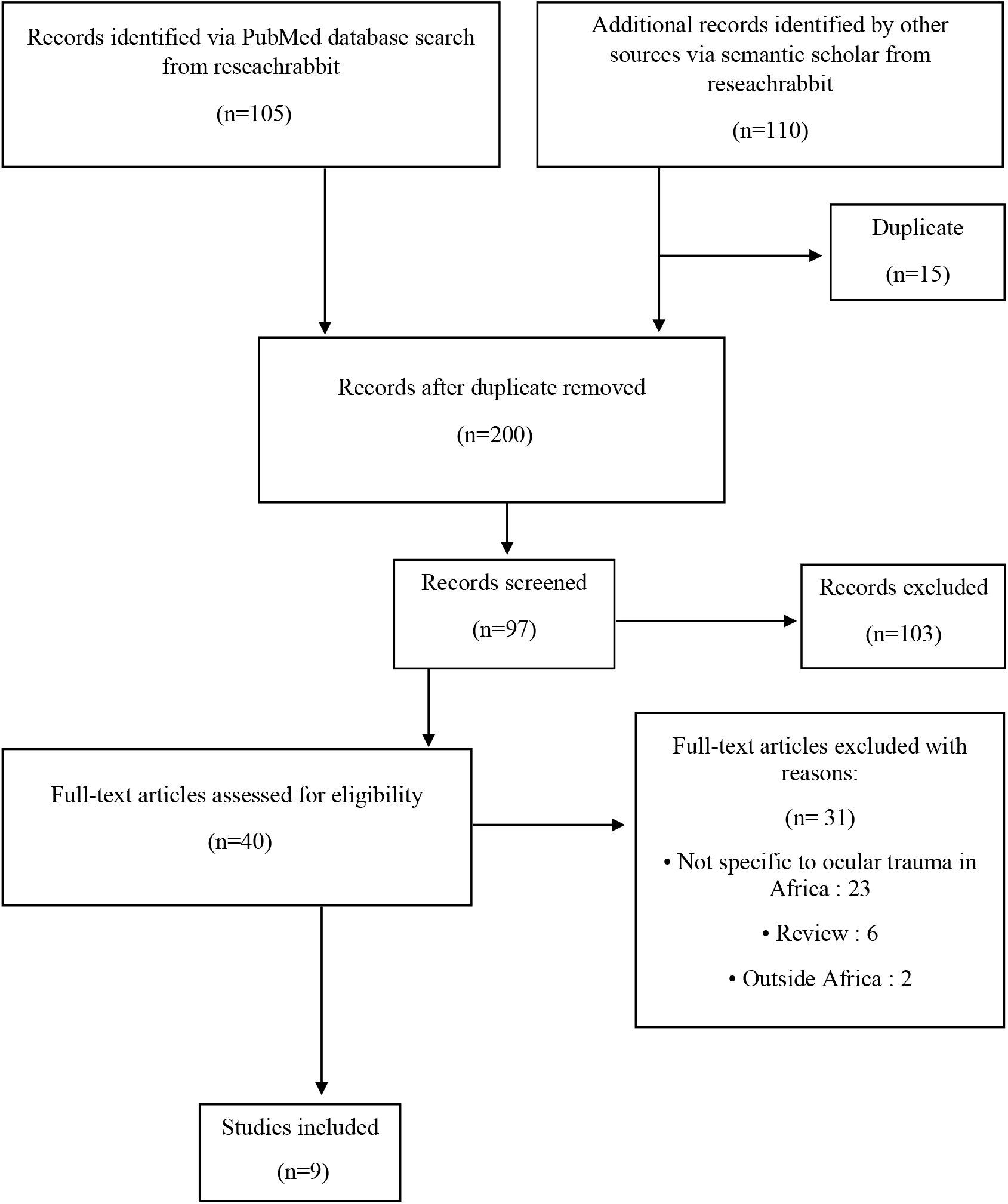

## Results

After screening, we obtained 40 full-text articles to evaluate for eligibility.

Articles that were excluded from these results: n= 31

- Not specific to eye trauma in Africa: 23
- Reviews: 6
- Outside Africa: 2

Nine articles were thus selected.

## COMMENTS ON THE RESULTS

**TABLE 1:** Frequency of eye trauma in Africa.

| FREQUENCIES | AUTHORS | COUNTRY | YEARS |
| --- | --- | --- | --- |
| 30.92% | PIERRE LOUIS LAMA et al. | GUINEA | 2022 |
| 19.5% | MELVIN CHETTY et al. | SOUTH AFRICA | 2022 |
| 3.2% | KELSY V. STUART et al. | SOUTH AFRICA | 2022 |
| 11% | C KRUSI, IL BRUCE et al. | SOUTH AFRICA | 2021 |
| 3.5% | CHINWE CYNTHIA et al. | NIGERIA | 2021 |
| 7.7% | DESTAW DAMTIE et al. | ETHIOPIA | 2020 |
| 3.4% | EMMANUEL K. ABU et al. | GHANA | 2020 |
| 44.1% | IRENE KIDA MINJA et al. | TANZANIA | 2019 |
| 35% | KERRY L. ALBERTO et al. | SOUTH AFRICA | 2019 |
| 30.7% | ONYINYECHUKWU MARY A. et al. | NIGERIA | 2019 |
| 3.1% | MICHAELINE A. and MODUPE A. | NIGERIA | 2017 |
| 19.6% | BOADI-KUSI SAMUEL B. et al. | GHANA | 2016 |
| 11.8% | ZALALENEM ADDISU MEHARI | ETHIOPIA | 2014 |

This table reports the frequency of eye trauma in Africa, reported from the year 2014, the highest frequency is observed in the studies IRENE KIDA MINJA et al. in 2019 **(39)** made in TANZANIA and the lowest frequency is observed in the studies of MICHAELINE A. and MODUPE A. in 2017 **(42)**, these data are not taken on a national scale although they were made in these aforementioned countries.

**TABLE 2:** Clinical aspect of ocular trauma in Africa.

| **CLINICAL SIGNS** | **COUNTRY** | **AUTHORS** | **YEARS** |
| --- | --- | --- | --- |
| Contusion (66.3%) | Guinea | PIERRE LOUIS LAMA et al. | 2022 |
| Closed globe injuries (72%) | Malawi | THOKOZANI ZUNGU et al. | 2021 |
| Penetrating injuries (60.8%) | Tunis | I. MALEK, L. NACEF et al. | 2020 |
| Eye injuries (39.5%) | Ghana | EMMANUEL K et al. | 2020 |
| Corneal tear (39.33%) | Ethiopia | KINDIE DESTA et al. | 2019 |
| Closed globe injury (41%) | South Africa | M. DJAN et al. | 2019 |
| Corneal scars (15.7%) | Eritrea | RAJENDRA GYAWALI et al. | 2017 |
| Traumatic cataract (12.06%) | Togo | KASSOULA B. et al. | 2016 |
| Corneoscleral laceration (67.9%) | Niger | CECILIA O. OJABO et al. | 2015 |
| Perforated globe from blunt force injury (25%) | Nigeria | CO ADEOTI and DS<br>AGBELEYE | 2015 |
| Corneoscleral rupture (63.2%) | Congo-Brazza | PW ATIPO-TSIBA | 2015 |
| Eye injuries (15.5%) | Ethiopia | BERAN SOLOMON et al. | 2014 |
| Open globe injury (26.4%) | Ethiopia | ALEMAYEHU WT and<br>SHAHIN S. | 2014 |
| Bruises (82%) | Benign | KOFI-MENSA SAVI et al. | 2013 |
| Contusion (32%) | South Africa | VN SUKATI and R<br>RAUTENBACH | 2013 |
| Open globe injury (81.3%) | Egypt | MAHA YOUSSEF et al. | 2013 |

This table illustrates the most common clinical signs. It appears that each study has its own particularity regarding the most common signs.

**TABLE 3:**
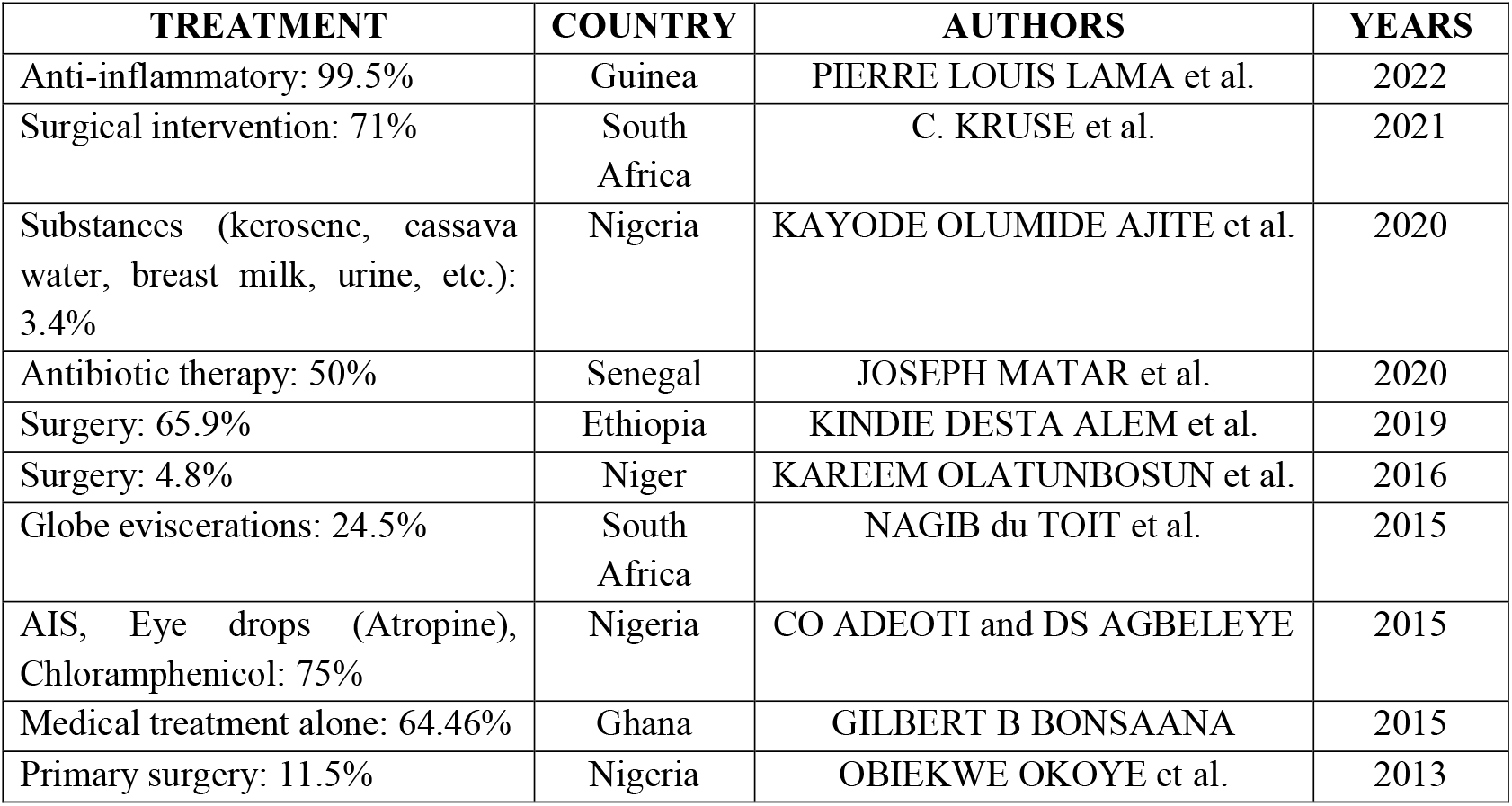
Therapeutic aspect of ocular trauma in Africa.

| **TREATMENT** | **COUNTRY** | **AUTHORS** | **YEARS** |
| --- | --- | --- | --- |
| Anti-inflammatory: 99.5% | Guinea | PIERRE LOUIS LAMA et al. | 2022 |
| Surgical intervention: 71% | South Africa | C. KRUSE et al. | 2021 |
| Substances (kerosene, cassava water, breast milk, urine, etc.): 3.4% | Nigeria | KAYODE OLUMIDE AJITE et al. | 2020 |
| Antibiotic therapy: 50% | Senegal | JOSEPH MATAR et al. | 2020 |
| Surgery: 65.9% | Ethiopia | KINDIE DESTA ALEM et al. | 2019 |
| Surgery: 4.8% | Niger | KAREEM OLATUNBOSUN et al. | 2016 |
| Globe eviscerations: 24.5% | South Africa | NAGIB du TOIT et al. | 2015 |
| AIS, Eye drops (Atropine), Chloramphenicol: 75% | Nigeria | CO ADEOTI and DS AGBELEYE | 2015 |
| Medical treatment alone: 64.46% | Ghana | GILBERT B BONSAANA | 2015 |
| Primary surgery: 11.5% | Nigeria | OBIEKWE OKOYE et al. | 2013 |

This table illustrates the most commonly encountered therapeutic aspects, resulting in each study having its own particularity on the most common therapy.

## Discussions

### The frequency of eye trauma

The frequency of eye injuries depends on the study environment and varies between regions and different age groups. However, other factors such as lifestyle, socioeconomic status, traffic conditions, sports and recreational activities and the type of data recording also influence this variability in frequency. **(16)**

The results showed an increasing trend of eye trauma. **(30)** The high frequency of eye trauma is due to the rural occupation with predominance of men (71.0%) than women due to the fact that in Africa men have preference in acquiring land for agriculture. **(43)** As confirmed by this study done in Ghana by Kindie Desta Alem et al 2019 **(47)**, 74% of eye injuries occurred in men because they are mainly engaged in outdoor activities where trauma is very common.

Most cases of ocular trauma were observed in the age group of 10 to 19 years and more than 75% of patients were under 40 years of age. **(15)** This is consistent with the findings of Nagib et al. **(62)**

In Africa, many children are cared for by elderly grandparents who often need care themselves. Children are therefore often left to fend for themselves and exposed to many dangerous objects such as sticks, bows and arrows, rubber ring missiles/brooms and wires. It is therefore not surprising that 51.0% of injuries occurred during play. **(51)**

Blunt objects were the most common cause of eye injuries followed by sharp wooden and metal objects together accounting for 27.2% however in Ethiopia, Tanzania and Nigeria a stick was the leading cause of eye injury. **(46)**

### Clinical manifestations

The signs found in the series included in this work vary from one country to another but in several studies **(65-67)** the clinical signs are dominated by open globe injuries representing 81.3% **(58)** and 18.7% of closed globe injuries. Among open globe injuries, 40% were corneal injuries, 6.6% were limbal injuries, 24% were scleral injuries, 27.8% were corneo-scleral injuries, 37% had iris prolapse, 20.5% had traumatic hyphema, 29.4% had traumatic cataract and 9.8% had an intraocular foreign body. **(58)**

### Management

The most commonly used management in our study was usually medical treatment **(68)** alone 64.46%, of which topical antibiotics alone/combined were the most common; 95.49% followed by analgesics alone/combined 40.09%. **(63)** This is consistent with a study in Ethiopia, where topical antibiotics and analgesics were the most common modality of treatment (63.4%). 35.54% eyes underwent surgical intervention of removal of foreign bodies from the ocular surface was the most common, 27.06%. **(55)**

## Conclusion

Ocular trauma, once described as a neglected disorder, has recently been highlighted as one of the major etiologies of monocular, noncongenital visual impairment and blindness in all regions of the world except Africa. Ocular trauma accounts for 7% of all bodily injuries and 10% to 15% of all ocular diseases. Particularly in developing countries, about half a million people are blind as a result of eye injuries and about 30-40% of cases of monocular blindness are due to eye trauma.

Etiopathological factors and their impact on visual health make it possible to highlight appropriate measures to reduce the number of eye injuries, which is imperative given the complications and serious after-effects leading to blindness. Achieving this objective requires prevention that involves institutions and communities.

## Data Availability

All data produced in the present work are contained in the manuscript

## Bibliographic references

1. M. Sissoko, N. Guirou, R. Romuld Elien GY, G. Saye, A. Simaga, H. Diallo et al. Eye trauma during the COVID-19 sanitary crisis at Iota-teaching hospital. University Hospital of the Tropical Ophthalmology Institute of Africa (IOTA University Hospital) / University of Science, Techniques and Technology of Bamako (USTTB), Medina Coura, Bamako, November 5, 2020.

2. M. Sidibe*, A. Dembele, A. Napo, O. Diallo, I. Conare, S. Fomba et al. Eye injuries by braid needle at the institute of tropical ophthalmology in Africa. SOAO Review - No. 01 – 2014, pp. 13–18.

3. P Louis*, Céougna S, Ibrahima F, Aly S, Sidikiba C, Balla Sovogui et al. Pediatric ocular trauma: Epidemiological, clinical and therapeutic aspects at CADESSO in Donka, Guinea, Open Journal of Ophthalmology, Vol.12, No.3, pp284–293, August 2022.

4. Lama P, C Sagno, I Fofana Sylla, S Camara, B Sovogui et al. Pediatric ocular trauma: Epidemiological, clinical and therapeutic aspects at CADESSO in Donka, Guinea, Open Journal of Ophthalmology, Vol.12, No.3, pp284–293, August 2022.

5. Y El Harrak, N Boutimzine and LO Cherkaoui. Occupational Eye Trauma: A Major Public Health Issue in Rabat, Morocco, European Journal of Public Health, Volume 29, Supplement number 4, November 2019, ckz186.329.

6. Miquele M, Grativol P, Natalia B and Fábio P. Epidemiological and occupational profile of ocular trauma in a reference center in Espírito Santo, Brazil; Revista Brasileira de Oftalmologia 76, 7–10, 2017.

7. A Kumar B, S Chigiri, Swathi M., Rohini M. and Shruthi T. Ocular trauma, International Journal of Research in Medical Sciences, December 2015, Vol 3, Issue 12, Page 3714–3719.

8. H Hashemi; Mehdi K; K Mohammad, and Akbar Fotouhi. History of Ocular Trauma in Tehran Population, Tehran Eye Study, Iranian Journal of Ophthalmology Volume 23, Number 3, 2011.

9. Rajendra P. M, Tanmay S, Virendra PS, CP Mishra, and Abdullah Al-Mujaini. Epidemiology of ocular trauma in northern India: a hospital-based study, Oman J Ophthalmol, 12(2): 78–83, 2019 May-Aug.

10. CC Jac-Okereke, Chukwunonso A, Regina E & Enujioke U. Current pattern of ocular trauma observed in tertiary institutions in South-Eastern Nigeria, BMC Ophthalmology Volume 21, Article No.: 420 (2021).

11. Hassan H; Mehdi K; Kazem M and A Fotouhi. History of Ocular Trauma in Tehran Population, Tehran Eye Study, Iranian Journal of Ophthalmology Volume 23, Number 3, 2011.

12. J. Ormeno I. Epidemiology and trends in hospitalizations for ocular trauma in Chile between 2001 and 2020, French Journal of Ophthalmology, Volume 45, number 9, November 2022, pages 1055–1062.

13. Sounouvou I, Zoumenou E. and Alamou S. Eye traumas in the emergency department of university hospital of Cotonou, July, 2014.

14. Kang Feng, Yi Yao, Zhi-Jun Wang, Hong-Ping Nie, Xiu Qin Pang, Hui Jin Chen et al. Mechanism and prognostic indicators of explosion-related ocular trauma: a study on vitrectomy of ocular injuries, Acta Ophtalmologica, Volume 1, Issue 1, 2016. 99, Issue 6, pp. e956-e962, January 8, 2021.

15. Chinwe C, C Jac-Okereke, I Ezegwui & RE Umeh. Current pattern of ocular trauma observed in tertiary institutions in South-Eastern Nigeria, BMC Ophthalmology Volume 21, Article No.: 420 (2021).

16. Dawood A. Ocular Trauma, Kashan University of Medical Sciences, Kashan, June 25, 2014.

17. J. Ormeno I. Epidemiology and trends in hospitalizations for ocular trauma in Chile between 2001 and 2020, French Journal of Ophthalmology, Volume 45, number 9, November 2022, pages 1055–1062.

18. Wanpeng W, Yalan Z, Jun Zeng, Meng S, Baihua Chen. Epidemiology and clinical characteristics of patients hospitalized for ocular trauma in South-Central China, Acta Ophthalmologica, Volume 95, Issue 6 p. e503–e510, March 30, 2017.

19. Huda S. Al-Mahdi MBBS, and Shakeel P. Hashim. Clinical pattern of pediatric ocular trauma in fast developing countries. Volume 19, Issue 4, October 2011, Pages 186–191.

20. Mustafa Iftikhar; Asad Latif; Ummara Z. Farid; Bushra Usmani; Joseph K. Canner; Syed MA Shah. Changes in the incidence of ocular trauma hospitalizations in the United States from 2001-2014. JAMA Ophthalmol. 2019 Jul;137(1):48-56.

21. Diane W and Allison R. Imaging of ocular trauma. June 27, 2022.

22. Miquele M, Patricia GC, Natalia B and Fábio P S. Epidemiological and occupational profile of ocular trauma in a reference center in Espírito Santo, Brazil; Revista Brasileira de Oftalmologia 76, 7–10, 2017.

23. Jungyul P, Sang C Yang, and Hee-young Choi. Epidemiology and clinical patterns of ocular trauma in a level 1 trauma center in Korea, 2021.

24. Keela Jing Xiea, Joshua Foremanab Hugh R., Taylorc Mohamed Dirania. The prevalence of vision loss due to ocular trauma in the Australian National Eye Health Survey, Injury Volume 48, Issue 11, November 2017, Pages 2466–2469

25. Kang Feng, Yi Yao, Zhi-Jun Wang, Hong-Ping Nie, Xiu Qin Pang, Hui Jin Chen et al. Mechanism and prognostic indicators of explosion-related ocular trauma: a study on vitrectomy of ocular injuries, Acta Ophtalmologica, Volume 99, Issue 6, pp. e956–e962, January 08, 2021.

26. M Brophy; SA. Sinclair; Sarah H et al. Pediatric Eye Injury–Related Hospitalizations in the United States. JUNE 01 2006.

27. S Martin-Prieto, C Álvarez-Peregrina, I Thuissard-Vassalo, C Catalina-Romero, Eva CB, C Villa-Collier and M Sánchez-Tena. Description of the epidemiology of eye lesions in an active population over 10 years old in Spain., Int. J. Approx. Res. Public Health 2020, 17 (12), 4454.

28. WA Tsiba. Management of eye trauma emergencies at the university teaching hospital of Brazzaville, Health Sci. Say: Vol 16 (1) January-February-March 2015.

29. Y El Harrak, N Boutimzine and LO Cherkaoui. Occupational Eye Trauma: A Major Public Health Issue in Rabat, Morocco, European Journal of Public Health, Volume 29, Supplement number 4, November 2019, ckz186.329.

30. Pierre Louis Lama*, Céougna Sagno, Ibrahima Fofana, Aly Sylla, Sidikiba Camara, Balla Sovogui et al. Pediatric ocular trauma: Epidemiological, clinical and therapeutic aspects at CADESSO in Donka, Guinea, Open Journal of Ophthalmology, Vol.12, No.3, pp284–293, August 2022.

31. Kaimbo Wa Kaimbo D, Spileers W and Missotten L. Eye emergencies in the Democratic Republic of Congo. BullSoc Belge Ophtalmol.2002:(284):49–53.

32. Pierre L, Aly S, Sidikiba C, Ceougna S, Oumar R, Ibrahima S. Incidence of primary open-angle glaucoma at the Flamboyants Communal Medical Center in Conakry, Guinea. Open Journal of Ophthalmology. Vol.12 No.3, August 2022.

33. Melvin CK, P. Simpie S. Occupational exposure to blood and body fluids among emergency medical service providers in the ethekwini metropolis of South Africa. African Journal of Emergency Medicine, Volume 12, Issue 2, June 2022, Pages 97–101

34. Kelsey V., Kian M, Robert N., Sharon Yl, Alasdair N., Mark C. Alcohol, intraocular pressure, and open-angle glaucoma. Ophthalmology. 2022 Jun;129(6):637–652.

35. C. Kruse, JL Bruce, W. Becker, DL Clarke. Management of ocular and periocular trauma should be coordinated according to ATLS principles and requires multidisciplinary collaboration. Injury. 2021 Sep;52(9):2606–2610.

36. Chinwe Cynthia Jac-Okereke, et al. “Current pattern of ocular trauma as seen in tertiary institutions in south-eastern Nigeria. » BMC Ophthalmology, vol. 21, no. 1, December 2021, 10.1186/s12886-021-02162-4.

37. D Damtie and Abraraw S. The prevalence of occupational injuries and associated risks among workers of Bahir Dar Textile Share Company, Amhara region, northwestern Ethiopia. Public Health J Environ. 2020 Jul 24;2020:2875297.

38. Emmanuel K, Stephen O, Jennifer A, Michael N, Enyam K. Epidemiology and visual outcomes of ocular injuries in a low-resource setting. Afr Health Sci. 2020 Jun;20(2):779–788. doi: 10.4314/ahs.v20i2.31.

39. Irene Kida Minja, et al. “Head and Neck Trauma in a Rapidly Growing African Metropolis: A Two-Year Audit of Hospital Admissions.” » International Journal of Environmental Research and Public Health, vol. 16, no. 24, December 2019, p. 4930, 10.3390/ijerph16244930.

40. Kerry LA, Trevor C, Aubrey M, Stephanus J. Canalicular lacerations: Causes, related ocular injury and management at St John Eye Hospital. African Vision and Eye Health 78 (1), 1–6, 2019.

41. O Angela-Mary, Aribabatemitayo P. Ocular Morbidity and Wearing of Protective Eyewear Among Carpenters in Mushin Musa Olatunbosun Kareem Local Government Area., 1Year: 2019 | Volume: 26 | Issue: 4 | Page: 199–204. Nigeria, Lagos.

42. Michaeline A Isawumi, et al. “Child Abuse and the Eye in an African Population. » Korean Journal of Ophthalmology, vol. 31, no. 2, March 2017, 10.3341/kjo.2017.31.2.143.

43. Boadi-Kusi Samuel Bert, et al. “Ocular injuries and eye care seeking patterns following injuries among cocoa farmers in Ghana. » African Health Sciences, vol. 16, no. 1, March 2016, p. 255 –65, 10.4314/ahs.v16i1.34.

44. Zelalem Mehari, and Zelalem Addisu Mehari. “Pattern of childhood ocular morbidity in rural eye hospital, Central Ethiopia”. BMC Ophthalmology, vol. 14, no. 1, April 2014, p. 50 –50, 10.1186/1471-2415-14-50.

45. Thokozani Zungu, et al. “Characteristics and visual outcome of ocular trauma patients at Queen Elizabeth Central Hospital in Malawi”. PLOS ONE, vol. 16, no.3, 2021, 10.1371/journal.pone.0246155.

46. Malek I, Sayadi J, Zerei N, Mekni M, El Amiria K, Zgolli H et al. Epidemiology and prognostic factors of open globe injuries in a Tunisian pediatric population, Fr J ophthalmol, vol 43, September 2020, 604–610.

47. Kindie Desta Alem, et al. “Profile of ocular trauma in patients presenting to the department of ophthalmology at Hawassa University: Retrospective study. » PLOS ONE, vol. 14, no. 3, March 2019, 10.1371/journal.pone.0213893.

48. M Djan, R Rautenbach. Hospitalized ophthalmic trauma in East London, South Africa. South African Journal of Ophthalmology 14(1), 21–26, 2019.

49. Rajendra G, Bharat K, Rabindra A, Arjun S, Rabindra P. Retrospective data on the causes of childhood visual impairment in Eritrea. BMC Ophthalmol. 2017. BMC Ophthalmol. 2017 Nov 22;17(1):209.

50. Kassoula B, Nidain M, Kokou V, Meba B, Ignace S, Kossi A et al. Management and functional outcomes of traumatic cataracts in the central region of Togo. The Pan African Medical Journal 25, 2016.

51. Cecilia O., Keziah N., and Olasupo S., Open globe injuries in Nigerians: epidemiological characteristics, etiological factors, and visual outcomes, Middle East Afr J Ophthalmol. 2015 Jan-Mar;22(1):69–73.

52. CO Adeoti, et al. “Banger-Related Ocular Injuries During New Year Festivities in Osogbo, SW Nigeria.” Ethiopian Journal of Health Sciences, vol. 25, no. 2, Apr. 2015, pp. 185– 88, 10.4314/ejhs.v25i2.12.

53. W Atipo T. Traumatic Eye Emergencies: Difficulties Related to Their Management at the Brazzaville University Hospital. HEALTH SCIENCES AND DISEASE 16 (1), 2015.

54. Berhan SD and Ephrem S D. Trends in eye diseases among children attending a tertiary university hospital: Southwestern Ethiopia. Ethiopia J Health Sci. 2014 Jan; 24(1):69–74.

55. Alemayehu W and Shahin S. Epidemiology of ocular injuries in Addis Ababa Ethiopia. J Ophthalmol East Cent South Africa. 2014; (July): 27–34.

56. KM Savi, Abel R, Patricia Y, Zakari N, Olivier B, Vicentia B. Contribution of ultrasound in ocular trauma in Parakou (Benin). Pan African Medical Journal 15 (1), 2013.

57. VN Sukati, et al. “Characteristics of eye injuries in urban KwaZulu-Natal Province, South Africa: 2005-2008”. African Vision and Eye Health, vol. 72, no. 3, July 2013, p. 119 –26, 10.4102/aveh.v72i3.285.

58. Maha Y, Magda A, Gihan H and Ahmed M. Epidemiological pattern of ocular trauma (retrospective study). Journal of the Egyptian Ophthalmological Society 106 (1), 42–42, 2013.

59. Kayode O, Iyiade A, Olusola J. Pattern of corneal disorders in Ekiti: a tertiary eye center experience. Annals of African Medicine 19 (2), 119, 2020.

60. Joseph M, Aboubacry S, Habsa K, Jean P, Aïssatou M, Aly M et al. Post-traumatic dehiscence on corneal graft at the Ophthalmological Clinic of Aristide Le Dantec Hospital Dakar. Pan Afr Med J. 2020; 37: 23.

61. Kareem O, Olufisayo T, Adeola O, Adekunle R, Folasade B. Indications for destructive ocular surgeries in a Nigerian tertiary eye care centre: a ten-year review. Nigerian Postgraduate Medical Journal 23(1), 12, 2016.

62. Nagib Du T, Hamza M and Colin C, Visual outcomes in patients with open globe injuries compared with predicted outcomes using the ocular trauma scoring system, Int J Ophthalmol. 2015;8(6):1229–1233.

63. Gilbert B. Review of ocular trauma in Tamale teaching hospital, H58/81148/2012 Ghana 2015.

64. Obiekwe O, Chimdi M, Nwabueze. Ten years of rural experience of surgical eye removal in a primary health care centre in south-eastern Nigeria. Rural and Remote Health 13 (2), 1–7, 2013.

65. Gain P. Ophthalm6/C2 exr.htm 12-05-09.

66. Ullern M., Roman S. Wounds and foreign bodies of the posterior segment. Encycl Med Chir (Elsevier, Paris), Ophthalmology, 21-700-A, 1999, 11p.

67. Hammami B., Feki J., Kamoun B., Ellouze S., Trigui A, Chaabouni M. Traumatic hyphema by contusion. About 40 cases. J.Fr. Ophthalmol., 1998, 21.10, 741–745.

68. Coulibaly My. Eye trauma in the ophthalmology department of Sikasso hospital: about 256 cases. Thesis Medicine 2018; FMPOS Bamako USTTB.

